# Death and new disability following bleeding complications during venoarterial extracorporeal membrane oxygenation. A prospective cohort study from the EXCEL registry

**DOI:** 10.1101/2025.06.16.25329735

**Authors:** Alastair Brown, Mark Dennis, Nivedita Rattan, Vinodh Bhagyalakshmi Nanjayya, Aidan Burrell, Ary Serpa Neto, Carol Hodgson, EXCEL Study Investigators and the International ECMO Network

**Author notes:** **Address for Correspondence and Reprints**: Prof Carol Hodgson.

## Abstract

**Introduction:** Bleeding events are common during the provision of Venoarterial ECMO (VA ECMO). Previous observational has suggested that these bleeding events are associated with an increased risk of in-hospital mortality however data on long term mortality and functional outcomes are lacking.

**Methods:** This multi-centre prospective observational study included patients enrolled in the EXCEL registry from February 2019 until June 2023. We collected baseline demographics, and details of ECMO support. The primary outcome was a composite of death or new disability (defined as an increase of ≥ 10%. in the world health organisation disability assessment schedule score) at 180 days after ECMO initiation. We used multivariable logistic and quantile regression adjusted for illness severity and patient characteristics and diagnosis to assess the association of bleeding with death and new disability and other functional and clinical outcomes.

**Results:** The final study cohort included 704 participants, median [interquartile range IQR] age 54.5 [42 to 64] years and 259 (36.8%) were female. Bleeding complications occurred in 312 (44.3%) participants and 154 (22.3%) had major bleeding. Patients with bleeding complications were more often receiving VA ECMO for peri-operative support [112 (36.4%) vs (73 (19.3%), p < 0.001). Patients who experienced bleeding complications were at significantly increased risk of the primary outcome at 6 months risk difference (RD) 8.95% (95% CI: 1.62 to 16.14). However, this finding was due to a significantly increased risk of new disability- adjusted RD 18.04% (95% CI: 6.22 to 29.55) rather than an increased risk of mortality in patients with bleeding compared to no bleeding [adjusted RD 5.71% (95% CI: -1.55 to 13.01)]. Patients with bleeding complications were also associated with an increased use of healthcare resources such as ICU length of stay and renal replacement therapy.

**Conclusion:** Bleeding increased the rate of death and disability at 6-months, but this was driven by increased disability rather mortality. Bleeding complications were associated with an increased use of healthcare resources. Prospective studies to address modifiable risk factors for bleeding complications are warranted.

## Introduction

The use of venoarterial extracorporeal membrane oxygenation (VA ECMO) for refractory cardiogenic shock has increased substantially in the last ten years^1^. Three recent meta-analyses have demonstrated that the use of VA ECMO is associated with an increased risk of vascular and bleeding complications compared to standard care ^2–4^. In addition, a Delphi study, including consumers and clinicians, identified bleeding complications as a core outcome for this support modality^5^.

Observational studies have previously demonstrated an association of bleeding complications during VA ECMO with increased mortality^6–9^, suggesting these complications may counteract the potential benefits of the therapy. Whilst there are many existing reports on types and rates of VA-ECMO complications, data on their effect on longer term functional outcomes post hospital discharge are very limited.^10,11^ Therefore, we conducted an analysis of an Australian and New Zealand ECMO registry, to assess the impact of bleeding complications on long-term survival, disability and quality of life in patients who have received VA-ECMO support.

## Materials and Methods

This was a prospective, multi-centre, registry-embedded cohort study of patients receiving VA ECMO in Australian and New Zealand Intensive care units (ICUs). The study was performed in accordance with the strengthening reporting of observational studies in epidemiology (STROBE) statement^12^.

All adult patients (≥18 years of age) receiving VA ECMO at participating sites between 18 February 2019 and 30 June 2022 were eligible for inclusion. Patients receiving VA ECMO assisted cardiopulmonary resuscitation (ECPR), defined as active chest compressions during cannulation, were excluded. For those with multiple VA ECMO runs, only the first run was included. Those patients who had a change in ECMO configuration during their run were included if their first run was on VA ECMO.

The EXCEL Registry is a bi-national registry, across 30 sites in Australia and New Zealand, that prospectively collect baseline, clinical and outcome data on all ECMO patients from participating institutions (Clinicaltrials.gov NCT03793257). Ethical approval was obtained prior to commencing the study from all sites, including a waiver of consent for hospital data and opt-out consent for 6-month follow-up (The Alfred HREC 534/18).

Data collection processes have been outlined in detail previously^13^. In brief, data were collected by trained research coordinators at participating sites and data monitoring was also conducted to ensure data quality. Baseline pre-ECMO data included demographics, cardiac arrest and diabetes status, frailty, and physiological parameters on the day of admission to ICU. Data were also collected for severity of illness and risk prediction scores, physiological and laboratory parameters, pharmacological interventions including sedation and vasoactive drugs. ECMO cannulation details, ECMO and mechanical ventilation settings, adjunctive therapies in ICU and adverse events were also collected. Death was confirmed in-hospital from the medical records by research coordinators or by telephone contact with next-of-kin at 6-months after ECMO initiation. Patients surviving the hospital admission were contacted by mail and phone. Patient-reported disability and health status were assessed using the 12-level World Health Organization Disability Assessment Schedule 2.0 (WHODAS) questionnaire and the EuroQoL 5 Dimensions – 5 Levels score (EQ5D-5L) at 6-months by telephone using trained central assessors. Data were collected from hospital admission until 6-months after ECMO initiation or death, whichever occurred first.

The primary outcome was a composite of death or new disability at 6 months post ECMO commencement measured at 180-days after ECMO commencement. New disability was defined as an increase ≥ 10% in the WHODAS^14^. Baseline disability was assessed at the 6 month follow up interview asking patients or carers to report the participants function prior to hospital admission. Other secondary outcomes included survival at hospital discharge and 6 months, hospital and ICU length of stay, use of renal replacement and disability at 6 months defined as a WHODAS of ≥25%,). Additional 6-month functional outcomes were quality of life measured by EuroQoL-Visual Analogue Scale (EuroQoL-VAS) and EQ-5D-5L. In addition, the EQ- 5D-5L utility score was calculated using UK population EQ-5D-5L health-status norms as at the time of analysis these were not available for Australia.

Bleeding complications were defined according to the site of bleeding and whether the bleeding event met the international society of thrombosis and haemostasis definition for major bleeding ^15^. Bleeding was determined according to review of the clinical records and laboratory results by the trained data collectors. Detailed definitions are available in the electronic supplementary material and are aligned with those of the international society of haematology and transfusion. Complication data were censored at ICU discharge or 7 days following ECMO discontinuation or death whichever occurred first. Patients who experienced multiple bleeding complications during ECMO were only counted once.

### Clinical Care

There is limited information within the registry data source regarding haemostatic and anticoagulation parameters. The majority of the sites within the registry follow one of three clinical guidelines^16–18^. These guidelines typically call for the administration of unfractionated heparin as first line with a target activated partial thromboplastin time (APTT) of 50-70 secs and a platelet count of > than 50-80,000. The recommended treatment during active bleeding included the cessation of heparin until 12 to 24 hours after bleeding had stopped and the judicious use of blood products (e.g. fresh frozen plasma, cryoprecipitate) and haemostatic drugs (tranexamic acid, protamine)^18^.

### Statistical Analysis

Continuous variables are reported as median and interquartile range (IQR) and categorical variables as number and percentage. To compare categorical variables, we used Fisher exact tests and to compare continuous variables we used Wilcoxon rank-sum tests (for 2 groups) or the Kruskal-Wallis test (for 3 or more groups). To estimate the effect of bleeding complications on the primary outcome and other categorial outcomes we fit generalised linear models with an identity link for univariate analyses and performed a linear mixed effect regression for adjusted analyses. Data on 180-day mortality, time to first complication and the time from first complication to death were compared between groups using a log rank test and are presented as Kaplan-Meier curves. All time intervals were taken from the point of ECMO initiation. When estimating the association of bleeding complications with death, patients with concomitant brain death were excluded. For continuous outcomes, we performed uni- and multivariable median regression using an interior point algorithm. For all adjusted models age, acute physiology and chronic health evaluation (APACHE) IV score, episode of cardiac arrest pre-ECMO, Charlson comorbidity score, use of renal replacement therapy before ECMO, lactate before ECMO and diagnosis were chosen *a priori* based on available evidence and clinical rationale. For the mixed effects regression model site was included as a random effect. No imputation was performed for missing data or outcomes. As bleeding complications were expected to occur in conjunction with neurological complications that have been reported to strongly associated with mortality^19–21^, we performed two sensitivity analyses. In the first we excluded patients with concomitant neurological complications and in the second we included the occurrence of a neurological complication in the mixed effects model.

## Results

Within the study period, 704 patients required VA ECMO support and had full complication data (eFigure 1); 259 (36.8%) were female; median age 54.5 years [IQR 42 to 64] and body mass index 27.2 [IQR 23.8 to 31.5]. The median SAVE and APACHE IV scores were -4.0 [IQR -8 to -1] and 79 [IQR 59 to 104], respectively, and the median duration of VA ECMO support was 4.9 days [IQR 2.9 to 7.9]. The most common indications for VA ECMO support were post cardiotomy 185 (27%) and acute myocardial infarction 155 (22.6%) (Table 1). Twenty-three (3.3%) patients required VA ECMO support for COVID-19 related myocardial dysfunction. The median lactate prior to VA ECMO initiation was 5.2 [IQR 2.5 to 9.1] and did not differ between those with and without bleeding. The majority of patients underwent percutaneous cannulation with 220 (29.4%) patients receiving surgical return cannulas and 142(19.0) receiving surgical access cannulas. One hundred and ninety patients had a cardiac arrest before ECMO (27.1%). Details of the ECMO support on first day of ECMO therapy are available in the electronic supplementary material (eTable 2).

**Figure 1:**
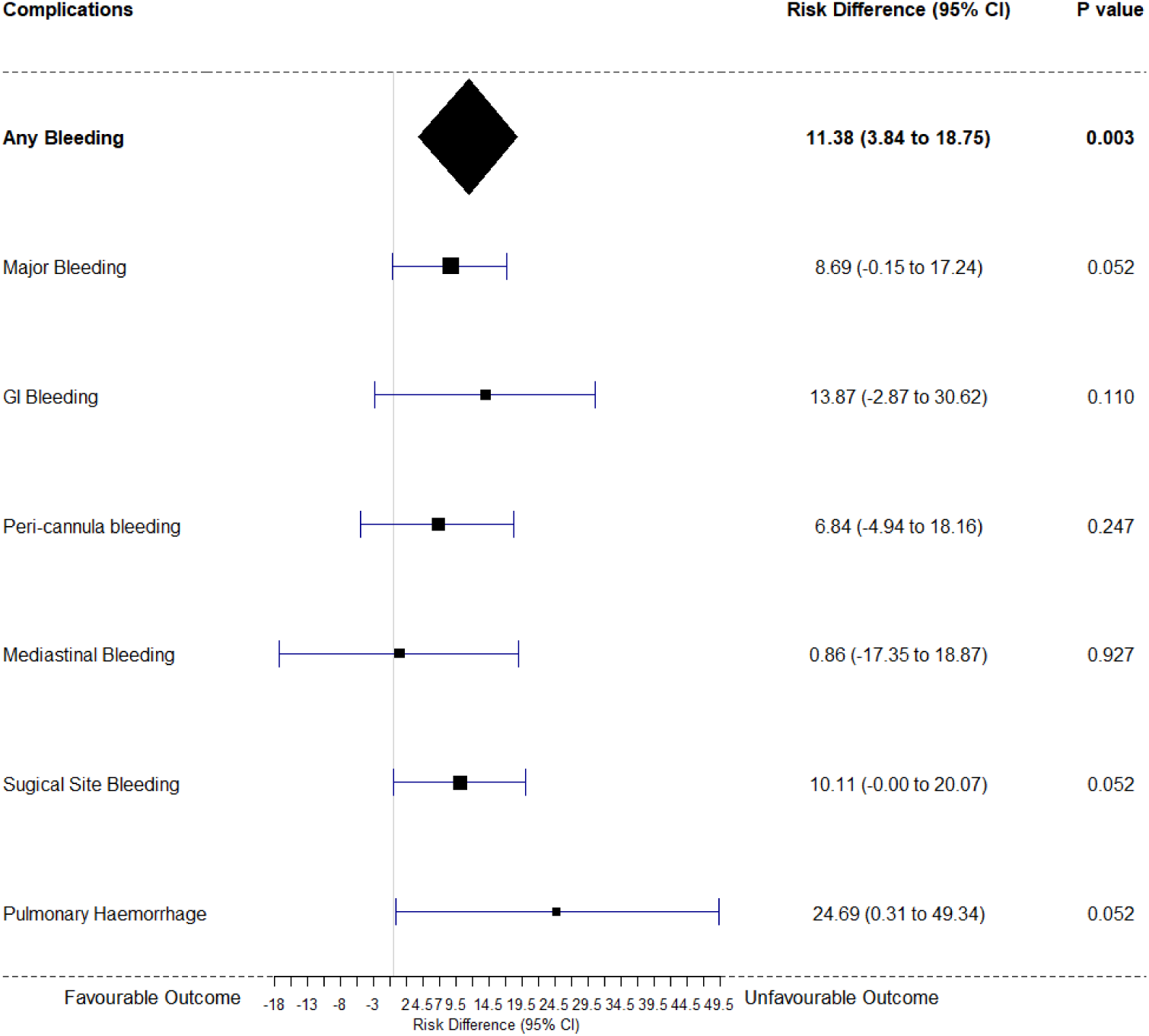
Forest Plot showing adjusted risk of death or new disability according to presence of bleeding complication and subgroups of bleeding complications.

**Table 1.**
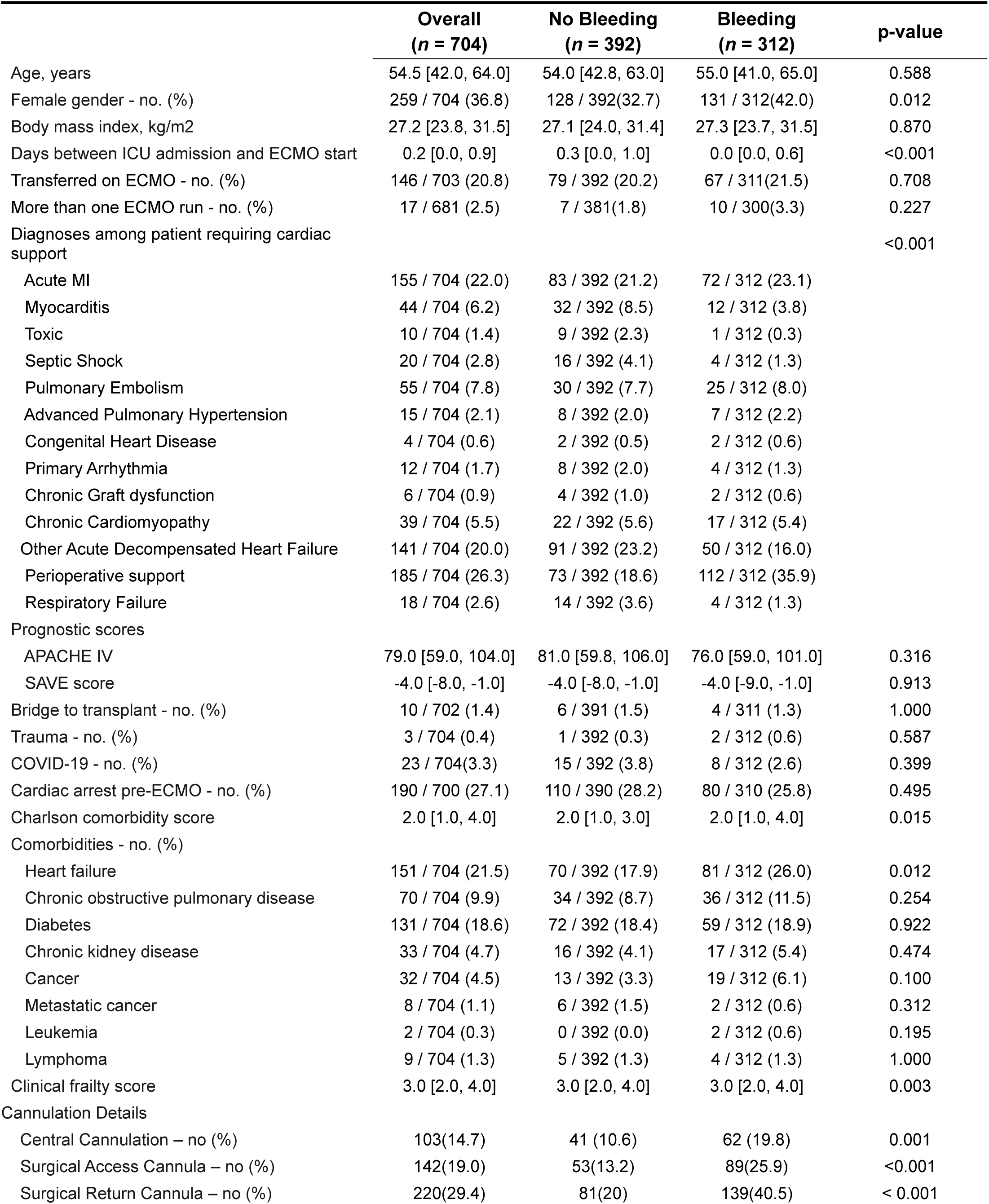
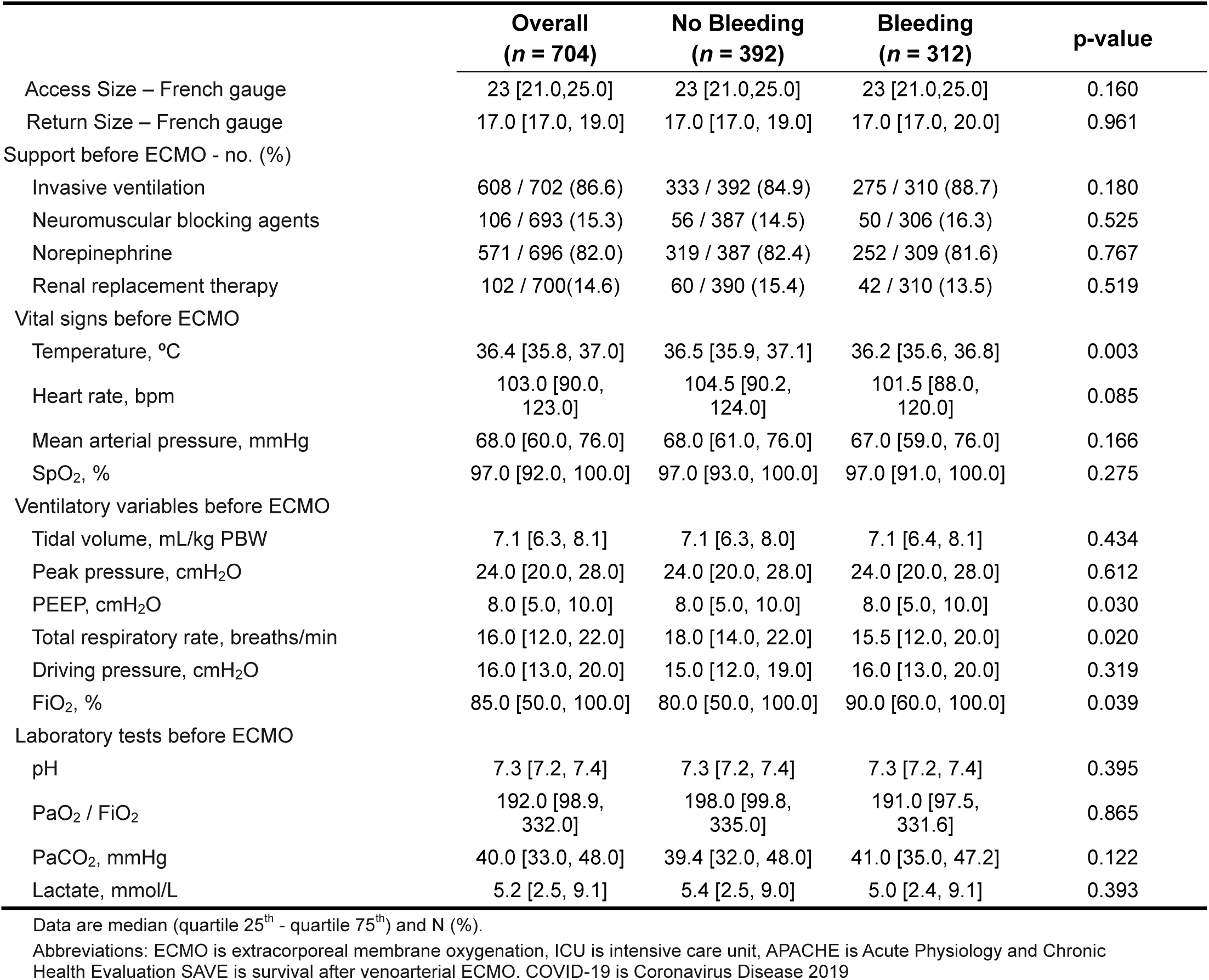
Baseline Characteristics.

|  | **Overall**<br>**(n = 704)** | **No Bleeding**<br>**(n = 392)** | **Bleeding**<br>**(n = 312)** | **p-value** |
| --- | --- | --- | --- | --- |
| Age, years | 54.5 [42.0, 64.0] | 54.0 [42.8, 63.0] | 55.0 [41.0, 65.0] | 0.588 |
| Female gender - no. (%) | 259 / 704 (36.8) | 128 / 392(32.7) | 131 / 312(42.0) | 0.012 |
| Body mass index, kg/m2 | 27.2 [23.8, 31.5] | 27.1 [24.0, 31.4] | 27.3 [23.7, 31.5] | 0.870 |
| Days between ICU admission and ECMO start | 0.2 [0.0, 0.9] | 0.3 [0.0, 1.0] | 0.0 [0.0, 0.6] | <0.001 |
| Transferred on ECMO - no. (%) | 146 / 703 (20.8) | 79 / 392 (20.2) | 67 / 311(21.5) | 0.708 |
| More than one ECMO run - no. (%) | 17 / 681 (2.5) | 7 / 381(1.8) | 10 / 300(3.3) | 0.227 |
| Diagnoses among patient requiring cardiac support |  |  |  | <0.001 |
| Acute MI | 155 / 704 (22.0) | 83 / 392 (21.2) | 72 / 312 (23.1) |  |
| Myocarditis | 44 / 704 (6.2) | 32 / 392 (8.5) | 12 / 312 (3.8) |  |
| Toxic | 10 / 704 (1.4) | 9 / 392 (2.3) | 1 / 312 (0.3) |  |
| Septic Shock | 20 / 704 (2.8) | 16 / 392 (4.1) | 4 / 312 (1.3) |  |
| Pulmonary Embolism | 55 / 704 (7.8) | 30 / 392 (7.7) | 25 / 312 (8.0) |  |
| Advanced Pulmonary Hypertension | 15 / 704 (2.1) | 8 / 392 (2.0) | 7 / 312 (2.2) |  |
| Congenital Heart Disease | 4 / 704 (0.6) | 2 / 392 (0.5) | 2 / 312 (0.6) |  |
| Primary Arrhythmia | 12 / 704 (1.7) | 8 / 392 (2.0) | 4 / 312 (1.3) |  |
| Chronic Graft dysfunction | 6 / 704 (0.9) | 4 / 392 (1.0) | 2 / 312 (0.6) |  |
| Chronic Cardiomyopathy | 39 / 704 (5.5) | 22 / 392 (5.6) | 17 / 312 (5.4) |  |
| Other Acute Decompensated Heart Failure | 141 / 704 (20.0) | 91 / 392 (23.2) | 50 / 312 (16.0) |  |
| Perioperative support | 185 / 704 (26.3) | 73 / 392 (18.6) | 112 / 312 (35.9) |  |
| Respiratory Failure | 18 / 704 (2.6) | 14 / 392 (3.6) | 4 / 312 (1.3) |  |
| Prognostic scores |  |  |  |  |
| APACHE IV | 79.0 [59.0, 104.0] | 81.0 [59.8, 106.0] | 76.0 [59.0, 101.0] | 0.316 |
| SAVE score | -4.0 [-8.0, -1.0] | -4.0 [-8.0, -1.0] | -4.0 [-9.0, -1.0] | 0.913 |
| Bridge to transplant - no. (%) | 10 / 702 (1.4) | 6 / 391 (1.5) | 4 / 311 (1.3) | 1.000 |
| Trauma - no. (%) | 3 / 704 (0.4) | 1 / 392 (0.3) | 2 / 312 (0.6) | 0.587 |
| COVID-19 - no. (%) | 23 / 704(3.3) | 15 / 392 (3.8) | 8 / 312 (2.6) | 0.399 |
| Cardiac arrest pre-ECMO - no. (%) | 190 / 700 (27.1) | 110 / 390 (28.2) | 80 / 310 (25.8) | 0.495 |
| Charlson comorbidity score | 2.0 [1.0, 4.0] | 2.0 [1.0, 3.0] | 2.0 [1.0, 4.0] | 0.015 |
| Comorbidities - no. (%) |  |  |  |  |
| Heart failure | 151 / 704 (21.5) | 70 / 392 (17.9) | 81 / 312 (26.0) | 0.012 |
| Chronic obstructive pulmonary disease | 70 / 704 (9.9) | 34 / 392 (8.7) | 36 / 312 (11.5) | 0.254 |
| Diabetes | 131 / 704 (18.6) | 72 / 392 (18.4) | 59 / 312 (18.9) | 0.922 |
| Chronic kidney disease | 33 / 704 (4.7) | 16 / 392 (4.1) | 17 / 312 (5.4) | 0.474 |
| Cancer | 32 / 704 (4.5) | 13 / 392 (3.3) | 19 / 312 (6.1) | 0.100 |
| Metastatic cancer | 8 / 704 (1.1) | 6 / 392 (1.5) | 2 / 312 (0.6) | 0.312 |
| Leukemia | 2 / 704 (0.3) | 0 / 392 (0.0) | 2 / 312 (0.6) | 0.195 |
| Lymphoma | 9 / 704 (1.3) | 5 / 392 (1.3) | 4 / 312 (1.3) | 1.000 |
| Clinical frailty score | 3.0 [2.0, 4.0] | 3.0 [2.0, 4.0] | 3.0 [2.0, 4.0] | 0.003 |
| Cannulation Details |  |  |  |  |
| Central Cannulation – no (%) | 103(14.7) | 41 (10.6) | 62 (19.8) | 0.001 |
| Surgical Access Cannula – no (%) | 142(19.0) | 53(13.2) | 89(25.9) | <0.001 |
| Surgical Return Cannula – no (%) | 220(29.4) | 81(20) | 139(40.5) | < 0.001 |

**Table 1 – Baseline Characteristics**
|  | **Overall**<br>**(n = 704)** | **No Bleeding**<br>**(n = 392)** | **Bleeding**<br>**(n = 312)** | **p-value** |
| --- | --- | --- | --- | --- |
| Access Size – French gauge | 23 [21.0,25.0] | 23 [21.0,25.0] | 23 [21.0,25.0] | 0.160 |
| Return Size – French gauge | 17.0 [17.0, 19.0] | 17.0 [17.0, 19.0] | 17.0 [17.0, 20.0] | 0.961 |
| Support before ECMO - no. (%) |  |  |  |  |
| Invasive ventilation | 608 / 702 (86.6) | 333 / 392 (84.9) | 275 / 310 (88.7) | 0.180 |
| Neuromuscular blocking agents | 106 / 693 (15.3) | 56 / 387 (14.5) | 50 / 306 (16.3) | 0.525 |
| Norepinephrine | 571 / 696 (82.0) | 319 / 387 (82.4) | 252 / 309 (81.6) | 0.767 |
| Renal replacement therapy | 102 / 700(14.6) | 60 / 390 (15.4) | 42 / 310 (13.5) | 0.519 |
| Vital signs before ECMO |  |  |  |  |
| Temperature, °C | 36.4 [35.8, 37.0] | 36.5 [35.9, 37.1] | 36.2 [35.6, 36.8] | 0.003 |
| Heart rate, bpm | 103.0 [90.0,<br>123.0] | 104.5 [90.2,<br>124.0] | 101.5 [88.0,<br>120.0] | 0.085 |
| Mean arterial pressure, mmHg | 68.0 [60.0, 76.0] | 68.0 [61.0, 76.0] | 67.0 [59.0, 76.0] | 0.166 |
| SpO<sub>2</sub>, % | 97.0 [92.0, 100.0] | 97.0 [93.0, 100.0] | 97.0 [91.0, 100.0] | 0.275 |
| Ventilatory variables before ECMO |  |  |  |  |
| Tidal volume, mL/kg PBW | 7.1 [6.3, 8.1] | 7.1 [6.3, 8.0] | 7.1 [6.4, 8.1] | 0.434 |
| Peak pressure, cmH<sub>2</sub>O | 24.0 [20.0, 28.0] | 24.0 [20.0, 28.0] | 24.0 [20.0, 28.0] | 0.612 |
| PEEP, cmH<sub>2</sub>O | 8.0 [5.0, 10.0] | 8.0 [5.0, 10.0] | 8.0 [5.0, 10.0] | 0.030 |
| Total respiratory rate, breaths/min | 16.0 [12.0, 22.0] | 18.0 [14.0, 22.0] | 15.5 [12.0, 20.0] | 0.020 |
| Driving pressure, cmH<sub>2</sub>O | 16.0 [13.0, 20.0] | 15.0 [12.0, 19.0] | 16.0 [13.0, 20.0] | 0.319 |
| FiO<sub>2</sub>, % | 85.0 [50.0, 100.0] | 80.0 [50.0, 100.0] | 90.0 [60.0, 100.0] | 0.039 |
| Laboratory tests before ECMO |  |  |  |  |
| pH | 7.3 [7.2, 7.4] | 7.3 [7.2, 7.4] | 7.3 [7.2, 7.4] | 0.395 |
| PaO<sub>2</sub> / FiO<sub>2</sub> | 192.0 [98.9,<br>332.0] | 198.0 [99.8,<br>335.0] | 191.0 [97.5,<br>331.6] | 0.865 |
| PaCO<sub>2</sub>, mmHg | 40.0 [33.0, 48.0] | 39.4 [32.0, 48.0] | 41.0 [35.0, 47.2] | 0.122 |
| Lactate, mmol/L | 5.2 [2.5, 9.1] | 5.4 [2.5, 9.0] | 5.0 [2.4, 9.1] | 0.393 |

Data are median (quartile 25<sup>th</sup> - quartile 75<sup>th</sup>) and N (%).
Abbreviations: ECMO is extracorporeal membrane oxygenation, ICU is intensive care unit, APACHE is Acute Physiology and Chronic Health Evaluation SAVE is survival after venaarterial ECMO. COVID-19 is Coronavirus Disease 2019

There were 312 patients receiving VA-ECMO (44.3%) who developed bleeding complications, of whom 154 (22.3%) had major bleeding. The median time to first bleeding event was 1.04 [IQR: 0.57 to 4.30] days. Surgical site bleeding was the most common location of bleeding 123 (17.5%) followed by cannulation site bleeding 82 (11.6%) while intracranial and pulmonary haemorrhage were the least common bleeding locations (eTable 3). Patients with bleeding complications were more often receiving ECMO for post cardiotomy (112 (36.4%) vs (73 (19.3%) and less often for Acute Decompensated Heart Failure (50 (16.2%) vs 91 (24.1%)) central cannulation was also more common among patients with bleeding (Table 1). Heparin was less often used on day 1 among bleeding patients (166 (53.2%) vs 287 (73.2%).

Primary outcome data was available for 607 (86.8%) of patients. Of these 422 of 607 (69.6%) patients had either died or had new disability at 6 months (including 308 patients (74.2%) who died and 114 (25.8%) patients who experienced new disability). Patients who experienced bleeding complications were at significantly increased risk of the primary outcome at 6 months RD 8.95% (95% CI: 1.62 to 16.14) The strength of the association increased further after adjusting for illness severity and treating centre (Table 3, Figure 2). However, the effect estimates were not consistent across both components of the primary outcome. Bleeding complications did not have significantly increased adjusted risk of mortality adjusted RD 5.71% (95% CI: -1.55 to 13.01) but did significantly increase the adjusted risk of new disability RD 18.04% (95% CI: 6.22 to 29.55). When considering the subgroups of bleeding complication the points estimates were all consistent with the main findings (Figure 1). The findings of the sensitivity analyses were consistent with the primary analysis (eTable 5 and 6).

**Figure 2:**
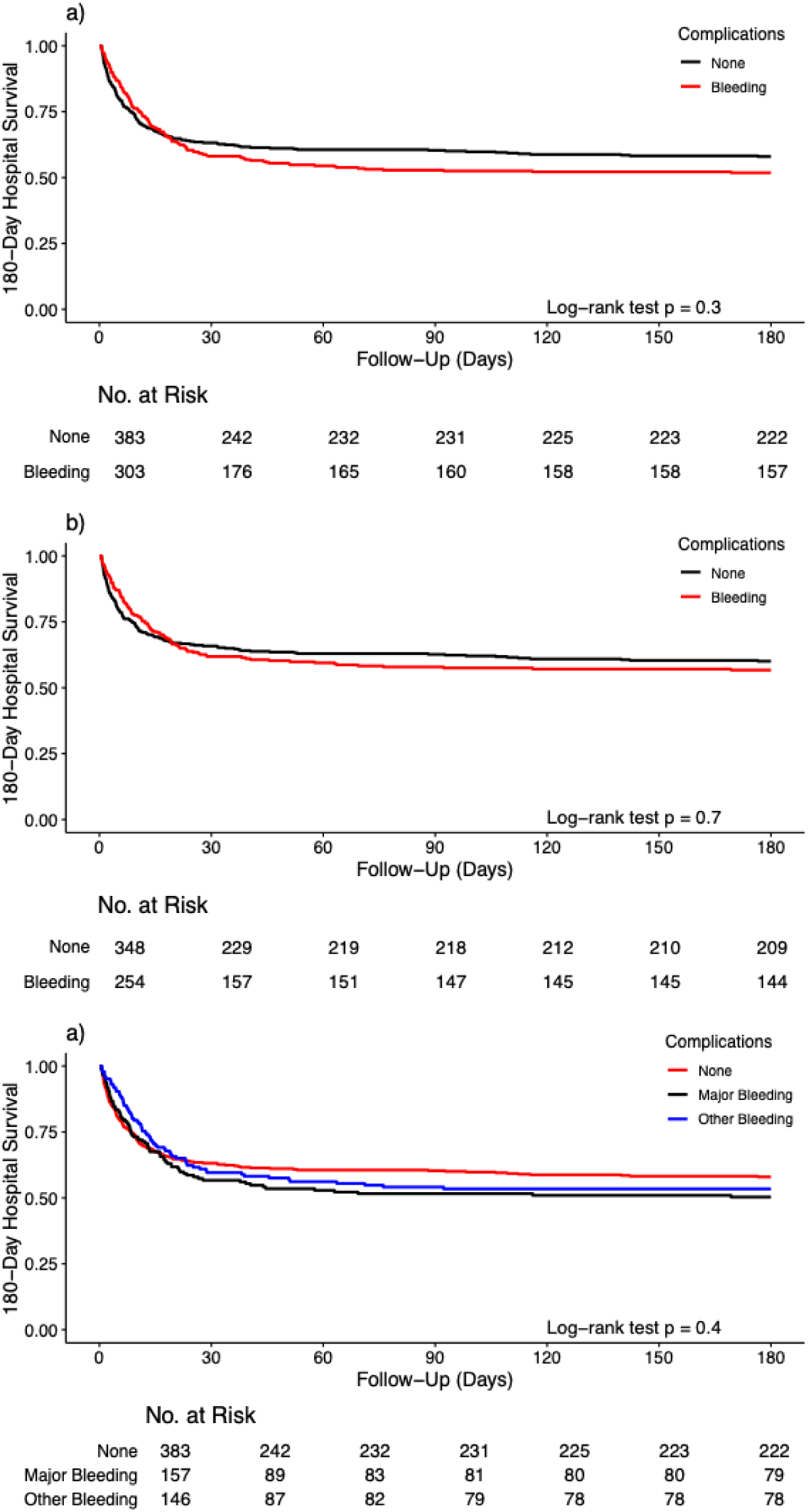
Kaplan-meier survival curves of survival to day 180 : a) Showing all results, b) Showing a sensitivity analysis excluding patients with neurological complications c) Comparison of survival curves between those with major bleeding and other forms of bleeding.

### Mortality at 6 months and complications

At 6 months, 146 (48.2%) of patients with a bleeding event had died compared to 162 (42.2%) of those without bleeding. There was non statistically significant increase in the risk of 6-month mortality for those experiencing a bleeding complication, after excluding patients with neurological complications the association remained insignificant and the survival curves appeared to converge(Figure 2a, b). There was also no significant difference in the rate of mortality between those with major bleeding and other bleeding complications (Figure 2c). Only 4 of the 704 patients (1.4%) died from hypovolaemic shock all of these were in the bleeding group (eTable 4).

### Disability and complications at 6 months

The rate of new disability was higher among patients with bleeding complications than those without these complications (43.9 vs 33.3%), although these differences were not statistically significant (Table 2). However, after adjustment for confounders the occurrence of bleeding complications was associated with a significantly increased risk of new disability RD, 18.04% (95% CI: 6.22 to 29.55). The median WHODAS score was also higher among patients with bleeding 16.7 [IQR: 7.8- 33.3] vs 12.5 [IQR: 4.2-29.2]. For each unit of blood transfused the mean WHODAS score increased by 0.3%, (eFigure 3). The proportion of participants with disability (WHODAS ≥ 25%) were similar among those with and without bleeding complications although the overall proportion of survivors with disability was high (33.8%), (Table 2). New disability seemed to be similar across all domains of the WHODAS (Figure 3).

**Figure 3:**
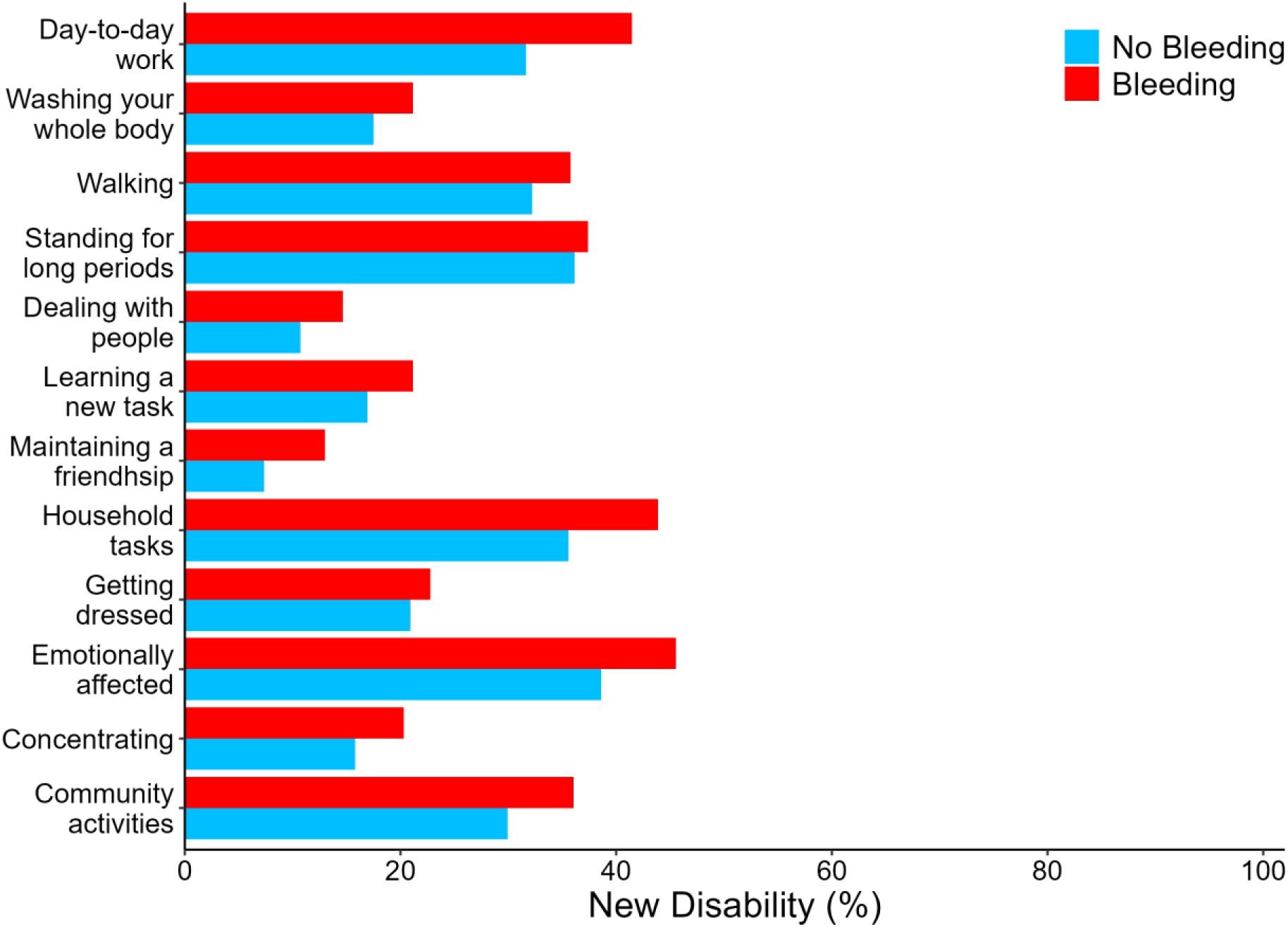
Comparison of domains of new disability between patients with and without bleeding complication

**Table 2:**
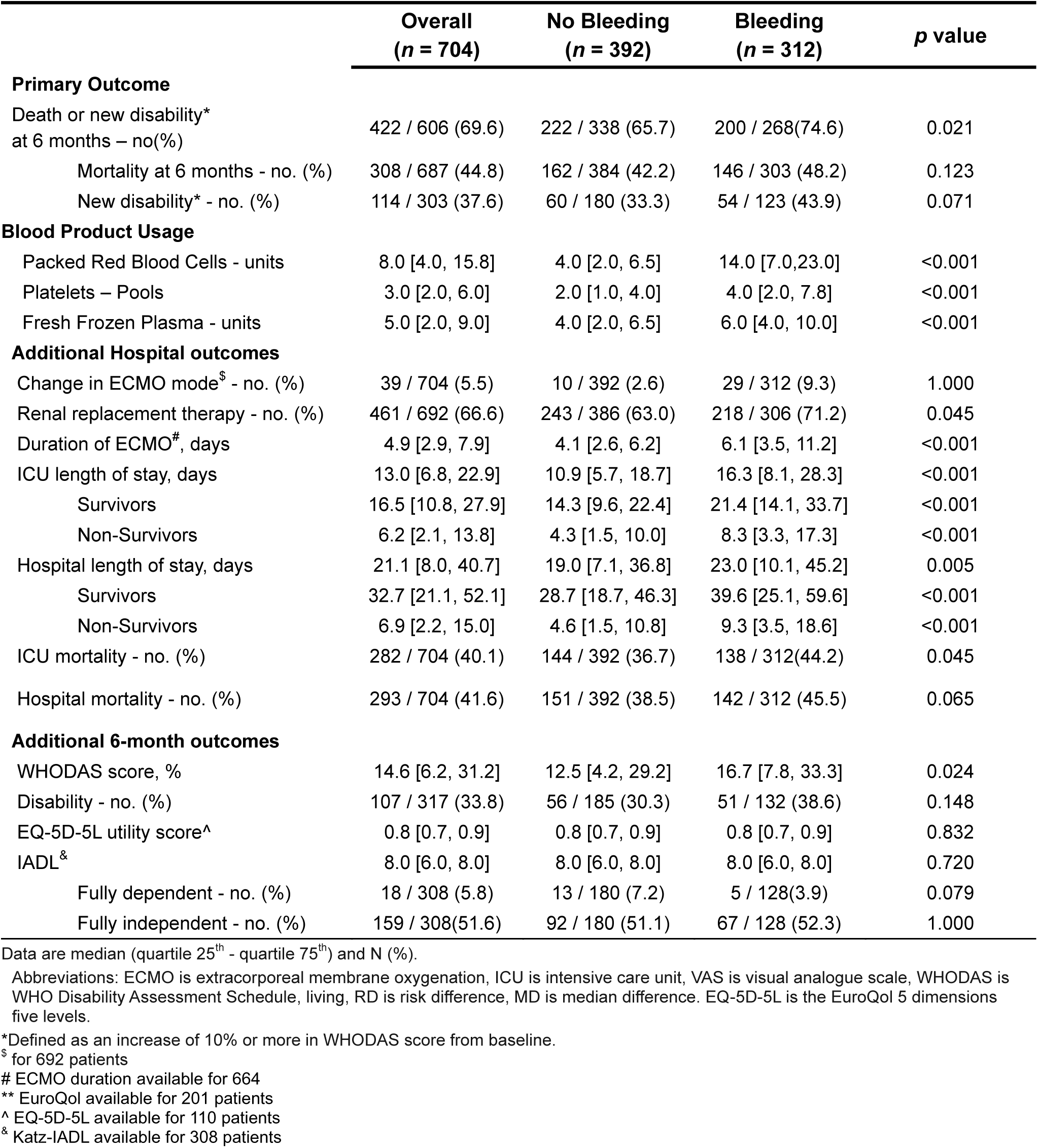
Outcomes.

### Additional functional outcomes at 6 months

There was no difference in health-related quality of life as measured by EQ-5D-5L between the groups or in the ability to perform instrumental activities of daily living. (Table 3).

**Table 3.** Adjusted and Unadjusted Outcomes – Entire Cohort.

| Primary Outcome and Components | No Bleeding<br>(*n* = 392) |  | Bleeding<br>(*n* = 312) |  |  |
| --- | --- | --- | --- | --- | --- |
|  |  | Unadjusted Effect Estimate<br>(95% CI) | *p* value | Adjusted Effect Estimate<br>(95% CI) | *p* value |
| Death on New Disability at 6 months | 1 (Reference) | RD 8.95 (1.62 to 16.14) | 0.016 | RD 11.81 (4.36 to 19.08) | 0.002 |
| Mortality at 6 months- no. (%) | 1 (Reference) | RD 6.00 (-1.49 to 13.46) | 0.116 | RD 5.71 (-1.55 to 13.01) | 0.131 |
| New Disability – no (%) | 1 (Reference) | RD 10.57 (-0.56 to 21.67) | 0.063 | RD 18.04 (6.22 to 29.55) | 0.003 |
| **Additional Hospital outcomes** |  |  |  |  |  |
| Change in ECMO mode - no. (%) | 1 (Reference) | RD 6.74 (3.33 to 10.56) | <0.001 | RD 6.79 (-0.31 to 13.94) | 0.066 |
| Renal replacement therapy - no. (%) | 1 (Reference) | RD 8.29 (1.25 to 15.23) | 0.020 | RD 5.87 (-0.92 to 12.87) | 0.097 |
| Duration of ECMO, days | 1 (Reference) | MD 1.94 (1.09 to 1.29) | <0.001 | MD 2.01 (0.79 to 3.24) | 0.001 |
| ICU length of stay, days | 1 (Reference) | MD 5.34 (3.24 to 3.18) | <0.001 | MD 4.58 (0.76 to 8.39) | 0.019 |
| Survivors | 1 (Reference) | MD 7.27 (3.48 to 5.73) | <0.001 | MD 7.08 (3.41 to 10.75) | <0.001 |
| Non-Survivors | 1 (Reference) | MD 3.96 (1.31 to 4.00) | 0.004 | MD 4.39 (1.86 to 6.92) | 0.001 |
| Hospital length of stay, days | 1 (Reference) | MD 3.98 (0.10 to 5.85) | 0.045 | MD 5.04 (-3.20 to 13.29) | 0.231 |
| Survivors | 1 (Reference) | MD 10.86 (5.26 to 8.47) | <0.001 | MD 9.54 (0.68 to 18.41) | 0.035 |
| Non-Survivors | 1 (Reference) | MD 4.61 (1.75 to 4.31) | 0.002 | MD 4.17 (1.43 to 6.90) | 0.003 |
| ICU mortality - no. (%) | 1 (Reference) | RD 7.50 (0.20 to 14.77) | 0.044 | RD 7.63 (0.63 to 14.75) | 0.037 |
| Hospital mortality - no. (%) | 1 (Reference) | RD 6.99 (-0.34 to 14.30) | 0.062 | RD 6.79 (-0.31 to 13.94) | 0.066 |
| **6-month outcomes** |  |  |  |  |  |
| WHODAS score, % | 1 (Reference) | MD 4.17 (-2.53 to 10.11) | 0.224 | MD 5.78 (3.15 to 8.42) | <0.001 |
| Disability - no. (%) | 1 (Reference) | RD 8.37 (-2.20 to 18.99) | 0.123 | RD 9.65 (-2.24 to 20.82) | 0.100 |
| EQ-5D-5L utility score | 1 (Reference) | MD -0.02 (-0.09 to 0.11) | 0.623 | MD -0.02 (-0.09 to 0.05) | 0.569 |
| IADL<sup>&</sup> | 1 (Reference) | MD -0.00 (-1.22 to 1.85) | 1.000 | MD 0.00 (-0.38 to 0.38) | 1.000 |
| Fully dependent - no. (%) | 1 (Reference) | RD -3.32 (-8.51 to 2.04) | 0.199 | RD - 1.59 (-7.48 tp 3.98) | 0.588 |
| Fully independent - no. (%) | 1 (Reference) | RD 1.23 (-10.07 to 12.50) | 0.831 | RD 2.79 (-9.38 to 14.93) | 0.663 |

Data are median (quartile 25<sup>th</sup> - quartile 75<sup>th</sup>) and N (%).
Models adjusted for: Age, Sex, Acute Physiology APACHE IV, cardiac arrest pre-ECMO, Charlson comorbidity score, renal replacement therapy, before ECMO, lactate before ECMO and diagnosis.
Abbreviations: ECMO is extracorporeal membrane oxygenation, ICU is intensive care unit, VAS is visual analogue scale, WHODAS is WHO Disability Assessment Schedule, living, RD is risk difference, MD is median difference. . EuroQol VAS is the EuroQol visual analogue scale. EQ-5D-5L is the EuroQol 5 dimensions five levels.

### Other hospital outcomes

The use of blood products was significantly higher among patients with bleeding complications (Table 2). The median length of stay in ICU for patients overall was 13.0 (IQR, 6.8 to 22.9) days. Among both survivors and non survivors the duration of ECMO, ICU and hospital length of stay were significantly longer and persisted after adjustment for confounders (Table 2).

## Discussion

This prospective cohort study demonstrated a significant association of bleeding complications with a composite outcome of death or new disability. This association was due to an increased risk of disability in patients with bleeding compared to no bleeding, and did not vary between patient with major bleeding and non-major bleeding. Patients with bleeding required significantly greater resources than patients without bleeding as measured by length of stay, use of renal replacement therapy and transfusion of blood products.

Our findings differ to the findings of previous investigations which have found an association of bleeding complications with an increased risk of hospital mortality^8–10,22^.. The distribution of bleeding sites was however similar to previous studies ^8^ although the rate of surgical site bleeding was slightly higher potentially due to a higher proportion of cardiac surgical patients in our cohort. Consistent with this heparin use was lower on day 1 in the bleeding group suggested clinicians withheld heparin in the face of active bleeding. Notably previous studies have included intracranial bleeding as a bleeding which we excluded from our cohort. and did not consider the concomitant occurrence of neurological complications in their analyses., It is therefore possible the effect of bleeding on mortality was overestimated in those previous studies, however notably the point estimate favoured the probability of increased mortality in those with bleeding albeit this was not statistically significant. Prior to this investigation there was very limited data assessing the association of bleeding with functional outcome in ECMO patients. Our finding suggest that bleeding is independently associated with an increased risk of worse functional recovery and with a significant increase utilisation of resources that have important clinical implications.

### Strengths and Limitations

Our study has several strengths as it provides the most comprehensive description of the association between bleeding complications and functional outcomes that has been presented. The data come from high quality, prospective observational study with clear and reproducible definitions and outcomes are assessed with validated tools. The follow-up rate is higher than reported in previous studies. Our analysis of the association of the dose of transfusion with increasing WHODAS score demonstrates a dose response of the need for transfusion with the risk of disability. Finally, by accounting for the presence of concomitant complications we were able to demonstrate the impact of bleeding on mortality was potentially smaller than previously described. We acknowledge limitations of our study. Firstly as an observational study, residual confounding cannot be excluded. Secondly data regarding patient’s coagulation status at baseline and throughout the ECMO run were not available, although the focus of our study was to assess the impact of complications rather than the risk factors of these complications. It is possible that our study was underpowered to detect a difference in mortality for patients with bleeding, and our point estimate suggested an increased risk of mortality. Finally, we did not have data on the dose of anticoagulation or target used for individual patients.

## Conclusion

Patients with bleeding complications during ECMO have increased risk of death and new disability at 6 months and increased use of clinical resources compared to patients without bleeding complications. This was primarily driven by increased disability rather than death. Prospective studies to address modifiable risk factors for bleeding complications are warranted.

## Data Availability

Data request made be to the EXCEL managment committee via the corresponding author

## Acknowledgments

We would like to thank the international ECMO network for their endorsement of this study.

